# A Randomized, Double-Blind Trial of the Analgesic and Anti-Inflammatory Effects of Naproxen Sodium and Acetaminophen Following Implant Placement Surgery

**DOI:** 10.1101/2022.12.30.22284065

**Authors:** Katherine N. Theken, Mengxiang Chen, D. Lucas Wall, Truongan Pham, Stacey A. Secreto, Thomas H. Yoo, Allison N. Rascon, Yu-Cheng Chang, Jonathan M. Korostoff, Claire H. Mitchell, Elliot V. Hersh

## Abstract

**Objectives:** The objectives of this study were to compare the analgesic and anti-inflammatory effects of naproxen sodium and acetaminophen after implant placement surgery.

**Materials and Methods:** Adult patients who received one or two dental implants were treated with naproxen sodium (440 mg loading dose + 220 mg q8h, n=15) or acetaminophen (1000 mg q6h - max daily dose 3000 mg, n=15) for three days after implant placement in a randomized, double-blind design. Pain was assessed on a 0-10 scale every 20 minutes for 6 h. Tramadol (50 mg) was available as a rescue medication. Plasma and gingival crevicular fluid (GCF) were collected prior to the surgery and 0, 1, 2, 4, 6, 24, and 72h after surgery for quantification of interleukin (IL)-6, IL-8, and IL-1β levels.

**Results:** Pain scores were significantly lower in patients treated with naproxen sodium compared to those treated with acetaminophen. Inflammatory mediator levels in plasma and GCF increased after surgery and returned to near baseline levels by 72h. Plasma IL-6 levels were significantly lower 6h after surgery in patients treated with naproxen sodium compared to acetaminophen. No differences in inflammatory mediator concentrations in GCF were observed between the treatment groups.

**Conclusions:** Naproxen sodium was more effective than acetaminophen in reducing post-operative pain and systemic inflammation following surgical placement of one or two dental implants. Further studies are needed to determine whether these findings are applicable to more complex implant cases and how they affect clinical outcomes following implant placement.

## Introduction

Placement of dental implants is a frequently performed outpatient surgical procedure, and the number of patients opting for this procedure over dentures and fixed bridges continues to increase (Elani et al., 2018). Implants have become the gold standard for replacing missing teeth due to their high level of predictability and patient acceptance, with long-term success rates greater than 95% (Ali & Kay, 2019; Schmitt & Zarb, 1993; Zarb & Schmitt, 1993). The soft tissue and bony trauma associated with dental implant surgery upregulates inflammatory mediators (Li et al., 2015; Pietruski et al., 2001), leading to post-operative pain that typically persists for several days after surgery (Al-Khabbaz et al., 2007; Alissa et al., 2009; Bockow et al., 2013; Hashem et al., 2006; Samieirad et al., 2017).

Numerous placebo-controlled studies support the use of non-steroidal anti-inflammatory drugs (NSAIDs) as first-line agents in managing pain following outpatient dental procedures due to their effectiveness and lack of addictive potential (Hersh et al., 2020; Moore & Hersh, 2013). Most of these studies have been performed in patients undergoing surgical extraction of bony impacted mandibular third molars, but similar results have been observed in studies of implant placement (Khouly et al., 2021; Mattos-Pereira et al., 2021; Melini et al., 2020). Pre-emptive administration of dexketoprofen 25 mg (Sanchez-Perez et al., 2018), ibuprofen 600 mg (Pereira et al., 2020), or piroxicam 40 mg (Bhutani et al., 2019), 15-60 minutes prior to surgery, resulted in less post-operative pain compared to placebo. However, dental implant patients are generally older with more comorbidities and concomitant medications than dental impaction patients (Aghaloo et al., 2019; Elani et al., 2018). Thus, there is greater concern for adverse events and drug interactions in this population. The pain intensity level following implant placement is generally less than dental impaction surgery (Al-Bayati et al., 2021; Al-Khabbaz et al., 2007), so over-the-counter (OTC) doses of NSAIDs and/or acetaminophen are an option for pain management in these patients. OTC dosing is more conservative than prescription dosing mainly for safety reasons (Hersh et al., 2007), and these lower dosages along with the shorter maximum durations of use (no more than 10-days) contribute to a side-effect profile no different from placebo (DeArmond et al., 1995; Kellstein et al., 1999). However, studies evaluating the comparative analgesic efficacy of OTC NSAIDs and acetaminophen in implant patients have not been performed.

Implant placement surgery also promotes a local and systemic inflammatory response. Elevated levels of interleukin (IL)-6, IL-8, and macrophage inflammatory protein (MIP)-1β have been observed in gingival crevicular fluid (GCF) from the implant site and the adjacent teeth one week after surgery and diminish over 12 weeks with subsequent healing (Emecen-Huja et al., 2013). Dental implant surgery also increases tumor necrosis factor (TNF)-*α* and IL-6 concentrations in plasma, indicative of a systemic inflammatory response (Li et al., 2015; Pietruski et al., 2001). Peak blood levels of these inflammatory mediators corresponded to peak pain intensity scores of 3-4 (mild to moderate pain) on a 0-10 visual analog scale in patients receiving an average of 2.5 implants (Li et al., 2015). However, the effects of OTC NSAIDs or acetaminophen on local or systemic levels of inflammatory mediators after implant placement have not been studied. Therefore, we sought to compare the analgesic and anti-inflammatory effects of OTC regimens of naproxen sodium and acetaminophen in patients following placement of one or two dental implants.

## Materials and Methods

### Study procedures

This was a randomized, double-blind, parallel-group pilot study in adult patients receiving one or two dental implants. Patients were recruited from the Graduate Periodontics and Penn Family Practice Clinics at the University of Pennsylvania School of Dental Medicine (PDM) enrolled between April 2021 and August 2022. Subjects having either maxillary or mandibular implant surgery, including those requiring bone grafting on the day of surgery, were included. The study protocol and informed consent document were reviewed and approved by the University of Pennsylvania Institutional Review Board (IRB protocol # 844440, ClinicalTrials.gov: NCT04694300). All patients provided informed consent prior to any study-related procedure.

Patients were excluded from the study if they presented with advanced periodontal disease (>20% Clinical Attachment Loss + >20% radiographic bone loss); had poor oral hygiene; smoked or used recreational drugs; were pregnant or nursing a child; had a history of bisphosphonate usage or systemic steroid use for longer than 2 weeks in the past 2 years; had a history of diabetes mellitus, substance use disorder, chronic pain, or inflammatory or autoimmune disease; or had contraindications to any of the study medications (naproxen sodium, acetaminophen, tramadol) including any scheduled or recent cardiac procedures (within 6 months), a history of asthma, urticaria or allergic-type/ hypersensitivity reactions after taking aspirin or other NSAIDs, a history of, or active gastrointestinal perforation, ulcer or bleeding, severe heart failure, anticoagulant use due to increased bleeding risk with concomitant naproxen sodium therapy, or antidepressant therapy due to risk of serotonin syndrome with concomitant tramadol use.

Baseline blood and GCF samples were collected prior to surgery. GCF samples were collected by inserting paper filter strips (Periopaper, Proflow, Amitville NY) into the gingival crevice of a tooth adjacent to the implant site (Aboyoussef et al., 1998). Surgical placement of one or two implants was performed according to standard of care in the PDM Graduate Periodontics and Penn Family Practice Clinics. Local anesthesia was achieved with lidocaine plus 1:100,000 epinephrine (1:50,000 for hemostasis when necessary), and/or 3% mepivacaine plain. Nitrous oxide sedation was allowed. The use of glucocorticoids or the long-acting local anesthetic 0.5% bupivacaine plus 1:200,000 epinephrine was not permitted.

Patients were randomized in a ratio of 1:1 to receive either naproxen sodium (440 mg loading dose followed by 220 mg q8h; maximum daily dose 660 mg) or acetaminophen (1000 mg q6h; maximum daily dose 3000 mg). These regimens were chosen to align with OTC dosing recommendations from the product labeling (*Aleve® Product Information*; *Tylenol® for health professionals*). The allocation sequence was generated by simple randomization, and the investigators and participants were blinded to allocation. The investigational pharmacist, who was not involved in the analysis of the study, maintained the unblinded allocation sequence and assigned participants to interventions sequentially. Drug blinding was performed by the University of Pennsylvania Investigational Drug Services (PENN IDS) employing an over-encapsulation technique.

After completion of the surgery, additional blood and GCF samples were collected (T=0), and patients received their first dose of blinded study medication (naproxen sodium 440 mg or acetaminophen 1000 mg) according to their randomization assignment. Patients then reported their pain intensity every 20 minutes throughout the 6-hour observation period using the 0-10 Numeric Rating Scale, where 0=no pain and 10=worst pain imaginable. Additional blood and GCF samples were collected at 1, 2, 4, and 6h post-study drug administration. Rescue analgesic (tramadol 50 mg) was allowed upon request. At the time of discharge, patients were asked to rate the effectiveness of the medication in managing their pain (completely or mostly effective, somewhat effective, somewhat ineffective, completely or mostly ineffective).

Following the 6-hour sample collection, patients were discharged with a blister pack containing blinded study medication, as well as rescue medication (tramadol 50 mg) in case of insufficient pain relief. Due to the differences in dosing interval between naproxen sodium and acetaminophen, the blister pack contained placebo capsules in addition to active medication to maintain blinding of treatment allocation (Table 1). Patients were given a diary to record outpatient analgesic use and pain intensity scores. They returned to clinic at 24 and 72 hours for blood and GCF collections, pill counts, and verification of pain diary entries. A global assessment of study medication (poor, fair, good, very good or excellent) occurred at 72 hours. This concluded patient participation in the study.

**Table 1:** Dosing schedule of study medication employed to maintain the blind

|  | Naproxen Sodium Group | Acetaminophen Group |
| --- | --- | --- |
| **Day One:** |  |  |
| Immediately post-surgery | 220 mg x 2 | 500 mg x 2 |
| 6 hours post-surgery | Placebo x 2 | 500 mg x 2 |
| 12 hours post-surgery | 220 mg x 1 plus placebo x 1 | 500 mg x 2 |
| **Day Two** |  |  |
| On Awakening | 220 mg x 1 plus placebo x 1 | 500 mg x 2 |
| 6 hours post dose one | Placebo x 2 | 500 mg x 2 |
| 8 hours post dose one | 220 mg x 1 plus placebo x 1 | Placebo x 2 |
| 12 hours post dose one | Placebo x 2 | 500 mg x 2 |
| 16 hours post dose one | 220 mg x 1 plus placebo x 1 | Placebo x 2 |
| **Day Three** |  |  |
| On Awakening | 220 mg x 1 plus placebo x 1 | 500 mg x 2 |
| 6 hours post dose one | Placebo x 2 | 500 mg x 2 |
| 8 hours post dose one | 220 mg x 1 plus placebo x 1 | Placebo x 2 |
| 12 hours post dose one | Placebo x 2 | 500 mg x 2 |
| 16 hours post dose one | 220 mg x 1 plus placebo x 1 | Placebo x 2 |

### Quantification of COX activity and inflammatory mediators

*C*yclooxygenase (COX)-1 activity was evaluated *ex vivo* by quantifying serum thromboxane B_2_ levels, as previously described (Patrignani et al., 1982). Briefly, whole blood was collected into vacuum tubes containing clot activator and incubated in a water bath at 37°C for 1 hour. Serum was separated by centrifugation and stored at -80°C until analysis. Thromboxane B_2_ levels were quantified using the Thromboxane B_2_ Express Monoclonal ELISA kit (Cayman Chemical, Ann Arbor, MI).

COX-2 activity was evaluated *ex vivo* by quantifying plasma prostaglandin (PG)E_2_ levels following lipopolysaccharide (LPS) stimulation in whole blood, as previously described (Panara et al., 1999). Briefly, heparinized whole blood was treated with aspirin (1mM) and incubated at room temperature for 15 minutes. LPS (*Escherichia coli*, serotype O111:B4, 10 µg/ml whole blood) was added, and the sample was incubated in a water bath at 37°C for 24 hours. Plasma was separated by centrifugation and stored at -80°C until analysis. PGE_2_ levels were quantified using the PGE_2_ Monoclonal ELISA kit (Cayman Chemical, Ann Arbor, MI).

Paper strips containing GCF were placed in 100 μl phosphate buffered saline (PBS), incubated at 4°C for 2 hours and stored at -80°C until analysis. IL-6, IL-8, and IL-1β levels in plasma and GCF were quantified using Human Quantikine ELISA kits (R&D Systems, Minneapolis, MN) according to the manufacturers’ instructions. Levels of IL-8 and IL-1β in plasma and IL-6 in GCF were below the limit of detection, so further statistical analyses were not performed.

### Statistical analysis

The primary endpoint for analgesic effects was a comparison of pain intensity scores between the naproxen sodium and acetaminophen groups. The primary endpoint for anti-inflammatory effects was a comparison of inflammatory mediator levels in plasma and GCF between the naproxen sodium and acetaminophen groups. A sample size of 30 patients (n=15/treatment) provided approximately 80% power to detect a 2-fold difference in these endpoints at *α*=0.05. Data are reported as mean ± standard deviation or median (interquartile range (IQR)). Baseline characteristics and biochemical measurements were compared by t-test or Mann-Whitney test, as appropriate. Pain scores and biochemical measurements over time were analyzed by mixed effect modeling, including time and treatment as main effects. All tests were two-sided and p<0.05 was considered statistically significant.

The CONSORT flow diagram and checklist are included in the Supplementary Information.

## Results

The study cohort included 30 adults (12 men and 18 women) with a mean age of 46.2±14.3 years. After implant placement surgery, 15 patients received naproxen sodium and 15 patients received acetaminophen. No significant differences in demographic or clinical factors were observed between the treatment groups (Table 2). All patients completed the inpatient sample collections and pain assessments. In the naproxen sodium group, one patient missed both follow-up visits and one patient missed the 72-hour follow-up. Both patients reported outpatient medication use and pain assessments, but global assessments were not provided.

**Table 2:** Demographic and clinical characteristics of the study population

|  | Naproxen sodium (n=15) | Acetaminophen (n=15) |
| --- | --- | --- |
| Men/Women | 8/7 | 4/11 |
| Age (years) | 46.8±14.9 | 45.5±14.2 |
| Body Mass Index (kg/m<sup>2</sup>) | 25.2±3.6 | 26.8±4.3 |
| Number of Implants |  |  |
| 1 implant | 12 | 12 |
| 2 implants | 3 | 3 |
| Bone Grafting |  |  |
| Yes | 1 | 2 |
| No | 14 | 13 |
| Length of Surgery (minutes) | 71.1±35.8 | 85.5±30.6 |

The study treatments were well-tolerated, with no serious adverse events observed throughout the study. During the inpatient period, one patient in the acetaminophen group reported agitation, nausea, and emesis following treatment with tramadol. During the outpatient period, one patient in the acetaminophen group reported an episode of emesis and one patient in the naproxen group reported a headache. All adverse events were mild and resolved without intervention.

### Pain assessments

Most patients reported no or mild pain after implant placement, with a median maximum pain score of 2 (IQR: 0-3.25) during the inpatient phase. Naproxen sodium was significantly more effective than acetaminophen in controlling pain during both the inpatient and outpatient periods (Figure 1). During the inpatient period, patients treated with acetaminophen reported a significantly greater maximum pain score (median: 3; IQR: 2-7) compared to patients who received naproxen (median: 1; IQR: 0-2; p<0.01). The inpatient effectiveness ratings and global evaluations (Table 3) favored naproxen sodium and were consistent with the pain intensity scores, but these differences were not statistically significant.

**Table 3:** Inpatient effectiveness rating and global evaluation

|  | Naproxen sodium (n=15) | Acetaminophen (n=15) |
| --- | --- | --- |
| Effectiveness Rating |  |  |
| Mostly or Completely Effective | 15 (100%) | 10 (66.7%) |
| Somewhat Effective | 0 (0%) | 2 (13.3%) |
| Somewhat Ineffective | 0 (0%) | 2 (13.3%) |
| Completely or Mostly Ineffective | 0 (0%) | 1 (6.7%) |
| Global Evaluation <sup>†</sup> |  |  |
| Poor | 0 (0%) | 0 (0%) |
| Fair | 0 (0%) | 1 (6.7%) |
| Good | 1 (7.7%) | 3 (20.0%) |
| Very Good | 4 (30.8%) | 5 (33.3%) |
| Excellent | 8 (61.5%) | 6 (40%) |

<sup>†</sup> Global evaluation was not collected for 2 naproxen-treated patients who did not attend the 72-hour follow-up visit

**Figure 1:**
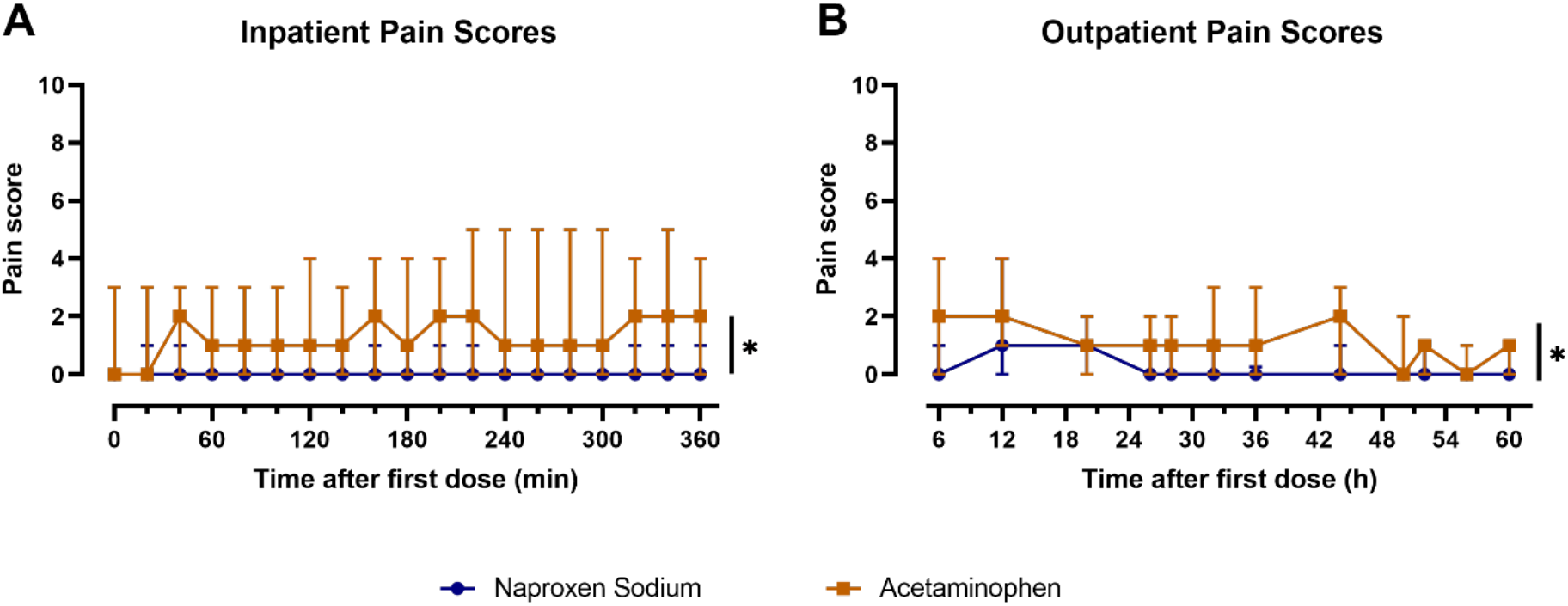
Comparison of median pain scores at each pain assessment during (A) inpatient and (B) outpatient periods between patients treated with naproxen sodium (blue) and acetaminophen (orange). Error bars indicate interquartile range (*p<0.05 for treatment).

Few patients required an opioid rescue analgesic. During the inpatient period, 3 patients in the acetaminophen group used tramadol, while no patients in the naproxen sodium group requested rescue analgesic. During the outpatient period, 5 patients in the acetaminophen group and 2 patients in the naproxen sodium group used tramadol. Of the patients who used tramadol during the outpatient period, 5 patients (2 naproxen sodium vs. 3 acetaminophen) used only one dose, 1 patient (acetaminophen) used 2 doses, and 1 patient (acetaminophen) used 3 doses.

### COX-1 and COX-2 inhibition

Naproxen sodium inhibited both COX-1 and COX-2 activity, while acetaminophen inhibited only COX-2 activity, as indicated by *ex vivo* whole blood assays (Figure 2). COX-1 activity *ex vivo* was inhibited by >90% in naproxen sodium-treated subjects during the inpatient period. Naproxen sodium tended to inhibit COX-2 activity *ex vivo* to a greater degree than acetaminophen, but this difference was not statistically significant (p=0.097 for treatment effect).

**Figure 2:**
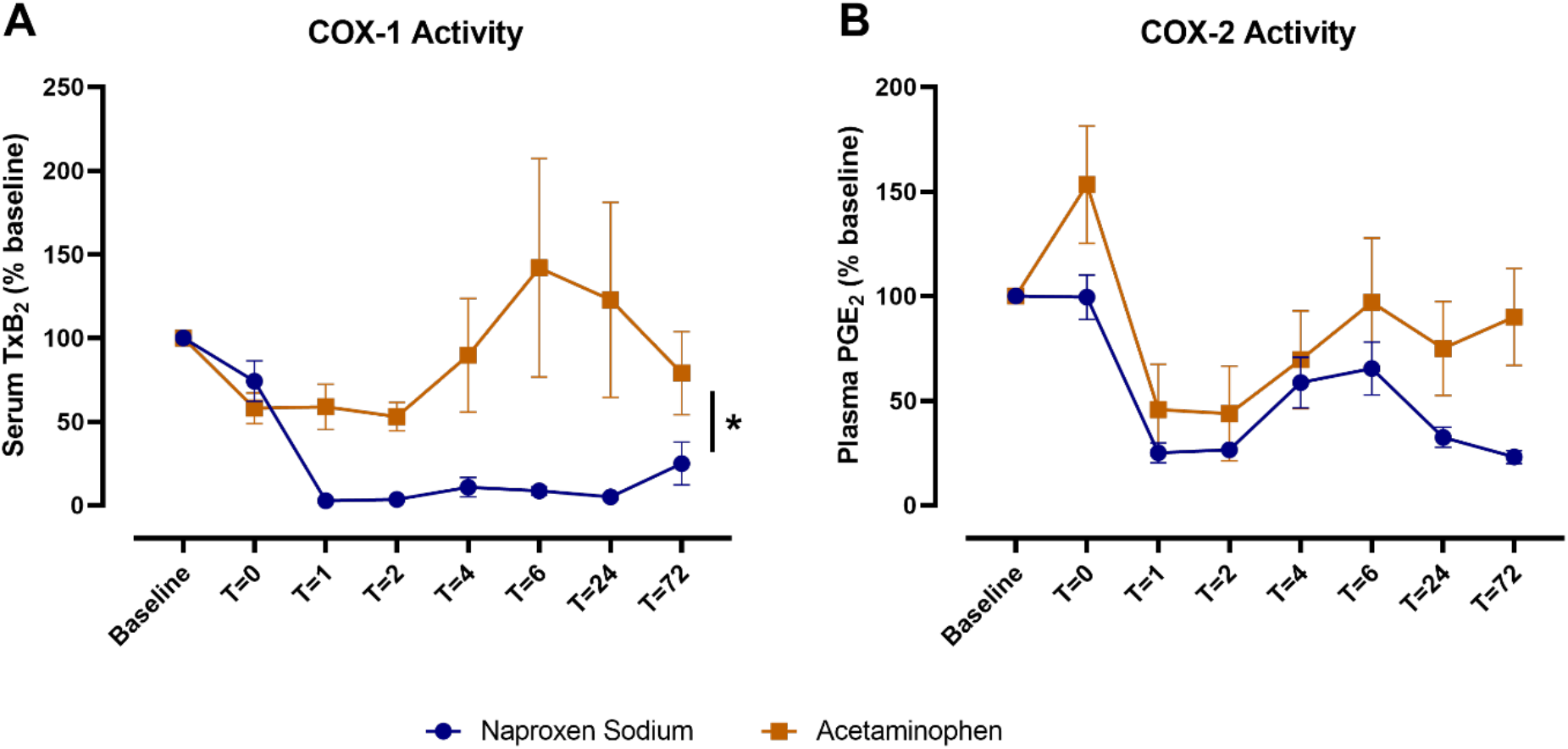
Comparison of (A) COX-1 and (B) COX-2 activity over time between patients treated with naproxen sodium (blue) and acetaminophen (orange). Data are expressed as a percent change from baseline and shown as mean ± SEM (*p<0.05 for time x treatment interaction).

### Systemic and local inflammatory mediator concentrations

Implant placement surgery increased plasma IL-6 concentrations, with the maximum levels observed 6 hours after surgery (Figure 3; p<0.05 for time effect). Naproxen sodium treatment blunted the increase in plasma IL-6 over time compared to acetaminophen (Figure 3A; p<0.05 for time x treatment interaction). At 6 hours, the median percent change in plasma IL-6 levels relative to baseline in patients treated with naproxen sodium was 276.9% (IQR: 181.3%- 348.4%) compared to 519.1% (IQR: 262.6%-1091%) in patients treated with acetaminophen (Figure 3B; p<0.05).

**Figure 3:**
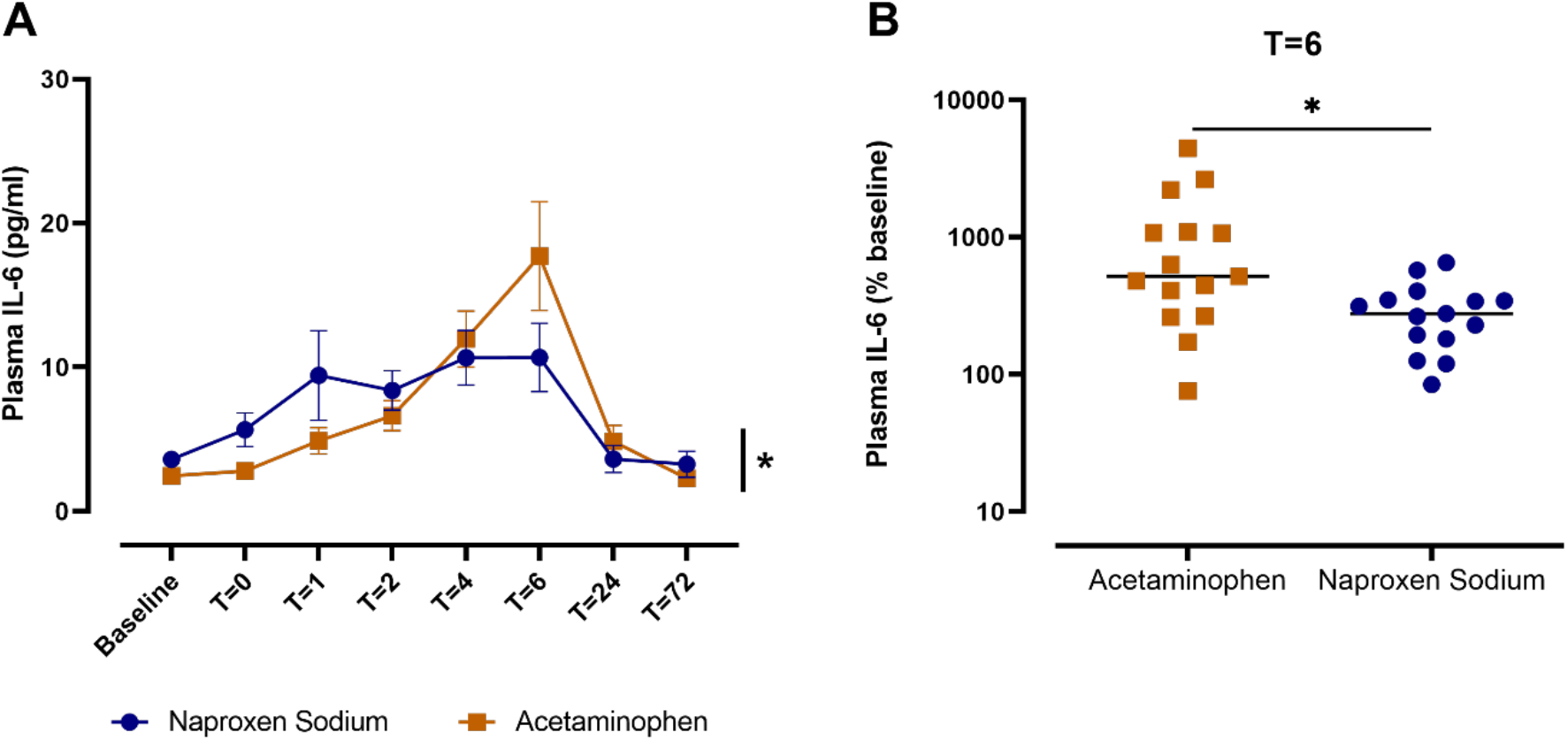
Comparison of plasma IL-6 levels between patients treated with naproxen sodium (blue) and acetaminophen (orange). (A) Plasma IL-6 concentrations over time by treatment. Data are shown as mean ± SEM (*p<0.05 for time x treatment interaction). (B) Plasma IL-6 levels at T=6 hr expressed as percent change from baseline. Crossbars indicate median, and data are plotted on a log_10_ scale (*p<0.05; Mann-Whitney test).

Implant placement surgery also increased IL-1β and IL-8 concentrations in GCF (Figure 4; p<0.05 for time effect). IL-1β levels peaked at 24 h and returned to near baseline levels by 72 h. IL-8 levels also increased over time, peaking at 24 h and remaining elevated at 72 h relative to baseline. No significant differences were observed between treatment groups for either IL-1β or IL-8 in GCF. After implant placement surgery, 70-85% of GCF samples were contaminated with blood during the inpatient period. For the 24 hr and 72 hr follow-up visits, approximately 40% of GCF samples were contaminated with blood.

**Figure 4:**
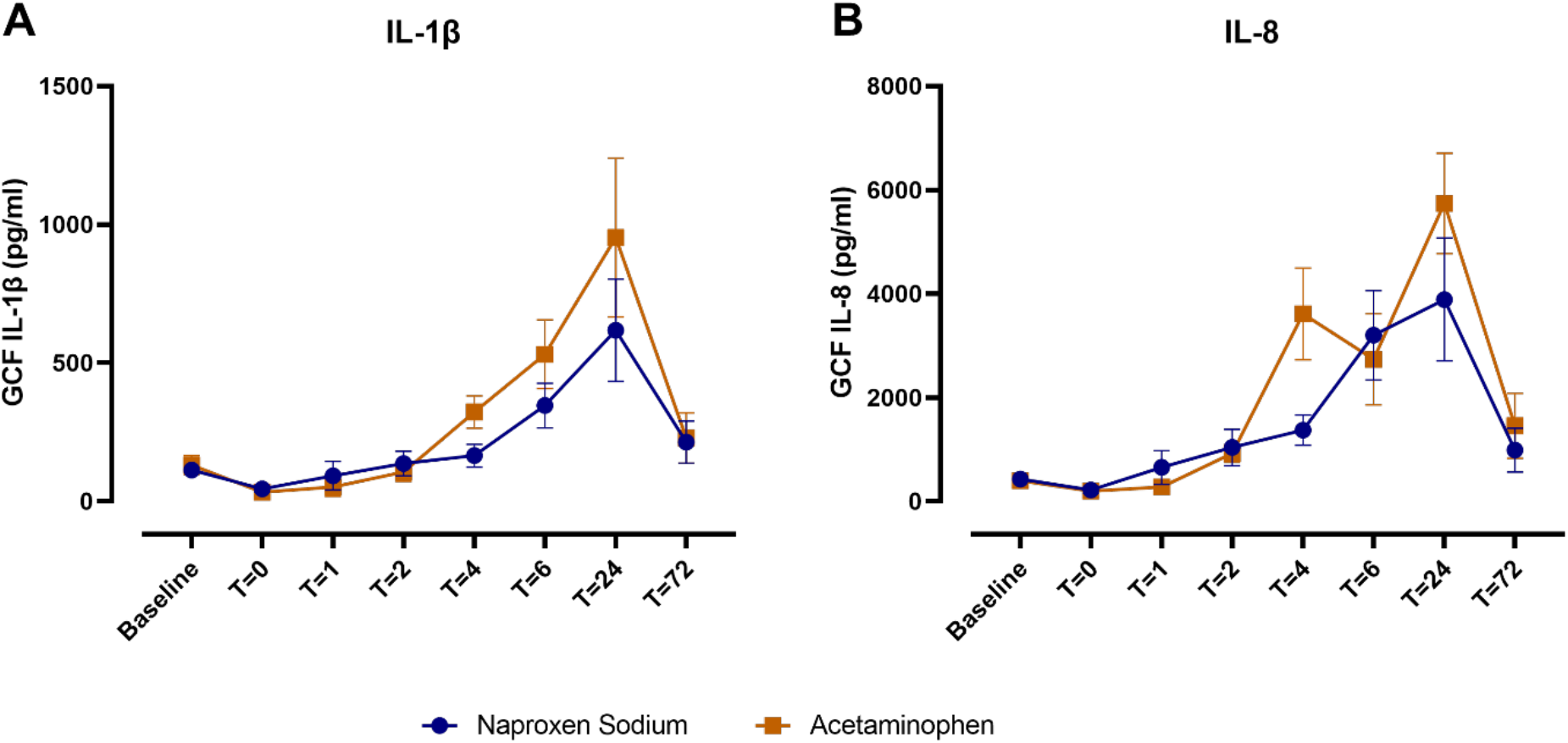
Comparison of (A) IL-1β and (B) IL-8 levels in gingival crevicular fluid (GCF) over time between patients treated with naproxen sodium (blue) and acetaminophen (orange). Data are shown as mean ± SEM.

We performed exploratory analyses to evaluate whether clinical or demographic factors influenced peak levels of inflammatory mediators in plasma (T=6 h) or GCF (T=24 h). Plasma IL-6 levels trended toward being higher 6 h after surgery in individuals who received two implants (n=6), compared to those who received one implant (n=24; p=0.10; Table 4). Naproxen sodium appeared to have an anti-inflammatory effect compared to acetaminophen in both groups (Figure 5), but the study was underpowered to formally evaluate this interaction. In contrast, the number of implants did not impact IL-1β or IL-8 levels in GCF at 24 h. Levels of both IL-1β and IL-8 in GCF at 24 h were significantly higher after implant placement in overweight and obese individuals (n=18; mean BMI: 28.6±2.1 kg/m^2^), compared to individuals in the healthy weight range (n=12; mean BMI: 22.0±2.5 kg/m^2^; Figure 6). No significant differences were observed between men and women for either plasma IL-6 or GCF IL-1β or IL-8 levels.

**Table 4:**
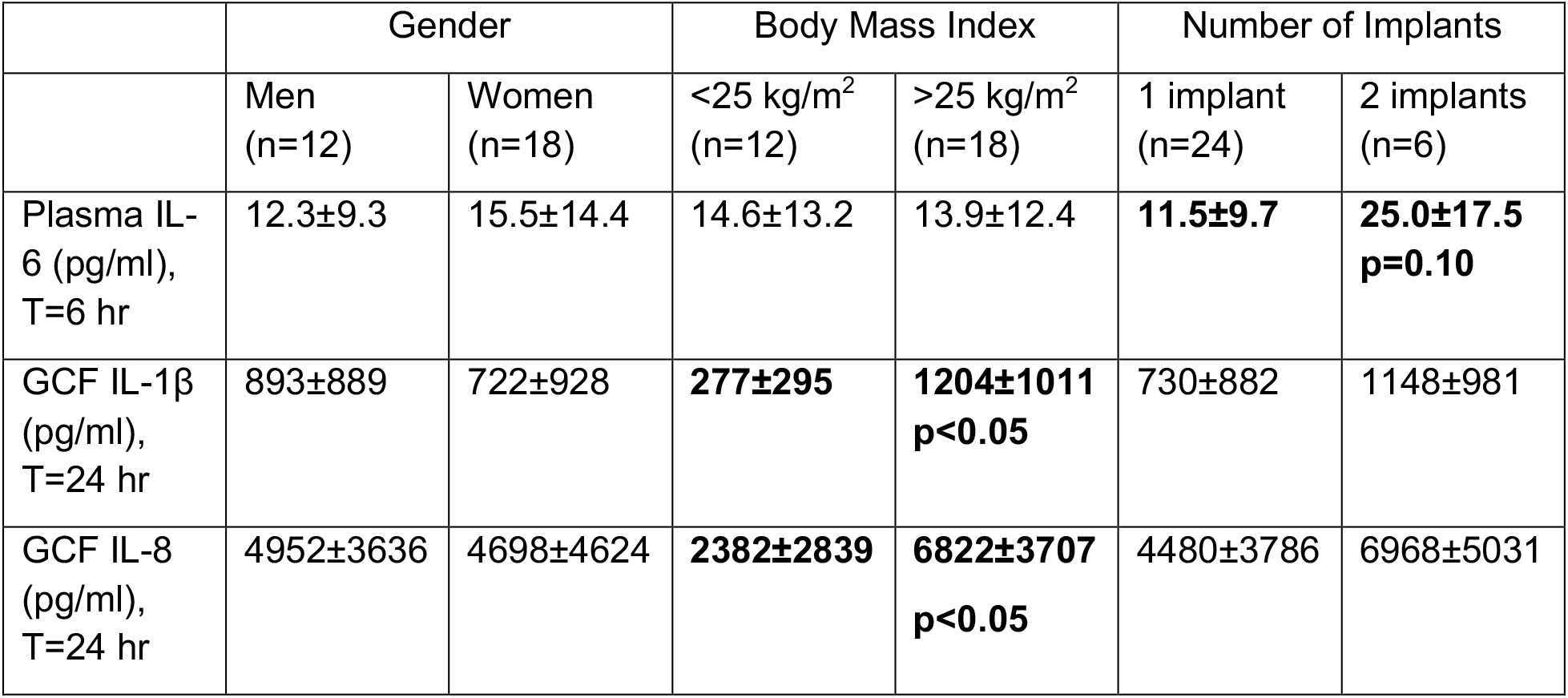
Peak inflammatory mediator concentrations by clinical and demographic characteristics

**Figure 5:**
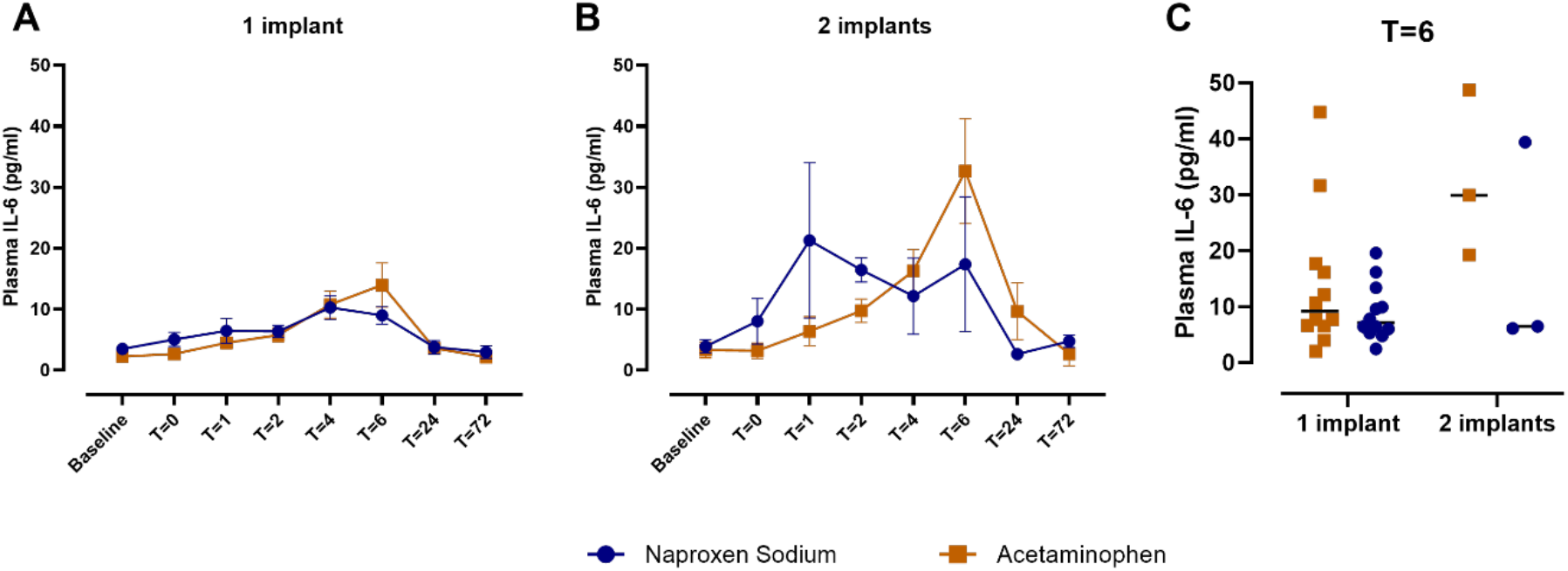
Comparison of plasma IL-6 levels between patients treated with naproxen sodium (blue) and acetaminophen (orange) by number of implants. (A) Plasma IL-6 concentrations over time in patients receiving one implant. Data are shown as mean ± SEM. (B) Plasma IL-6 concentrations over time in patients receiving two implants. Data are shown as mean ± SEM. (C) Plasma IL-6 concentrations at T=6 h by treatment and number of implants. Crossbars indicate median.

**Figure 6:**
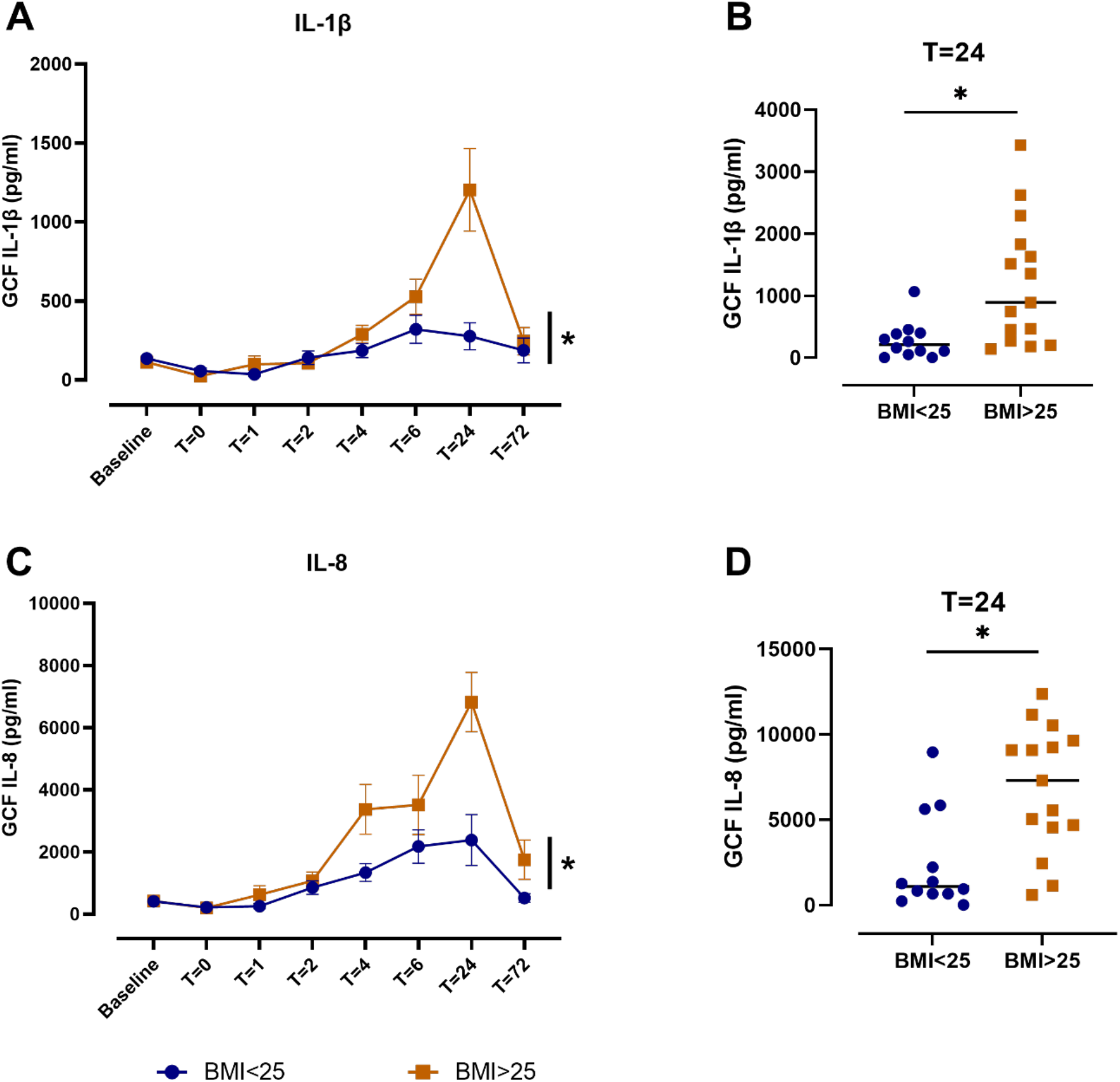
Comparison of IL-1β and IL-8 levels in gingival crevicular fluid (GCF) between patients with body mass index (BMI)<25 kg/m^2^ (blue) and BMI>25 kg/m^2^ (orange). (A) IL-1β concentrations in GCF over time. Data are shown as mean ± SEM (*p<0.05 for BMI). (B) IL-1β concentrations in GCF at T=24 h. Crossbars indicate median (*p<0.05; Mann-Whitney test). (C) IL-8 concentrations in GCF over time. Data are shown as mean ± SEM (*p<0.05 for BMI). (D) IL-8 concentrations in GCF at T=24 h. Crossbars indicate median (*p<0.05; Mann-Whitney test).

## Discussion

Nonaddictive analgesics, including NSAIDs and acetaminophen, are recommended as first-line agents in the management of pain following outpatient dental procedures (Hersh et al., 2020). Prior studies have demonstrated that prescription doses of NSAIDs are superior to placebo in reducing post-operative pain and swelling following implant placement surgery (Khouly et al., 2021; Mattos-Pereira et al., 2021; Melini et al., 2020). However, to our knowledge, this is the first study to compare the analgesic and anti-inflammatory effects of OTC doses of an NSAID and acetaminophen following implant placement surgery.

We observed that naproxen sodium was more effective than acetaminophen at reducing post-operative pain, despite most patients reporting only mild pain intensity. This is consistent with the results of prior studies performed in patients following third molar extraction. A single dose of naproxen sodium 440 mg was superior to acetaminophen 1000 mg in peak analgesic effects and duration of action (Kiersch et al., 1994). Naproxen sodium 440 mg also produced analgesic effects at least equivalent to and for some endpoints superior to optimal doses of acetaminophen 650 mg plus hydrocodone 10 mg following third molar extraction (Cooper et al., 2021).

Implant placement surgery promoted a systemic inflammatory response in the acute post-surgical period, as indicated by induction of IL-6 in plasma. Similarly, Pietruski and colleagues observed that serum concentrations of IL-6 and IL-8, but not IL-1, increased the day after implant placement compared to baseline in a small cohort (N=10) (Pietruski et al., 2001). Another study reported increases in plasma TNF-α and IL-6 concentrations 4 h after implant placement, which were significantly lower in patients who received dexmedetomidine during surgery compared to those who received midazolam (Li et al., 2015). We observed that patients treated with naproxen sodium had lower plasma IL-6 levels compared to acetaminophen-treated patients 6 h after surgery, consistent with a systemic anti-inflammatory effect of NSAID treatment.

We also observed induction of IL-1β and IL-8 in GCF following implant placement surgery, which peaked at 24 h after surgery. Few studies have measured inflammatory mediator concentrations in GCF in the acute post-surgical period. Blood contamination was present in the majority of GCF samples collected after surgery. However, neither IL-1β nor IL-8 was detected in plasma; thus, it is likely that the level of blood contamination had minimal effects on our results. Interestingly, we observed no differences in either IL-1β or IL-8 levels in GCF between the treatment groups. Rather, the levels of these inflammatory mediators differed based on body weight. Notably, these effects were apparent only after surgery, and no differences in IL-1β and IL-8 levels in GCF between healthy weight and overweight/obese patients were observed at baseline. One study in men who had an implant for at least 12 months reported that obese patients had higher peri-implant bleeding on probing, peri-implant probing depth, peri-implant marginal bone loss and levels of whole salivary IL-1β and IL-6, compared to patients with a healthy weight. However, all obese patients had a history of periodontitis, compared to no patients in the control group, so these observations may be confounded by disease status (Abduljabbar et al., 2016). Future studies will be necessary to elucidate the mechanisms underlying the elevated levels of IL-1β and IL-8 in GCF after surgery in overweight and obese patients and determine the effect on clinical outcomes.

There are limitations to our study. This was a pilot study with only 30 participants. Thus, our analysis was not powered to comprehensively evaluate the clinical and demographic factors that influence the local and systemic inflammatory response to implant placement surgery or drug response. In addition, we excluded smokers and patients with diabetes, autoimmune diseases, or other comorbidities that might influence the inflammatory response to implant placement surgery. Although this limits potential confounding, it precludes interrogation of the influence of these factors on response to naproxen sodium or acetaminophen. The post-surgical GCF samples were taken from teeth adjacent to the surgical site, and the sulci may have been damaged due to the incision. Therefore, we cannot ensure that the samples were truly GCF, rather than an inflammatory exudate. Finally, we followed patients for only 72 h after surgery and did not collect data regarding clinical outcomes like peri-implantitis or implant failure.

In conclusion, our study demonstrates that naproxen sodium is more effective than acetaminophen in reducing post-operative pain and systemic inflammation following surgical placement of one or two dental implants. These findings lay a foundation for future studies to evaluate how clinical and demographic factors impact the response to analgesic therapy following implant placement surgery.

## Supporting information

CONSORT checklist

CONSORT flow diagram

## Data Availability

All data produced in the present study are available upon reasonable request to the authors.

## Acknowledgements

This study was supported by an investigator-initiated research grant from Health Care LLC. The funder had no role in the design and conduct of the study; collection, management, analysis, and interpretation of the data; preparation, review, or approval of the manuscript; and decision to submit the manuscript for publication.

