## Supplementary figures and images for "A Randomized, Double-Blind Trial of the Analgesic and Anti-Inflammatory Effects of Naproxen Sodium and Acetaminophen Following Implant Placement Surgery"

### CONSORT flow diagram

#### CONSORT Flow Diagram

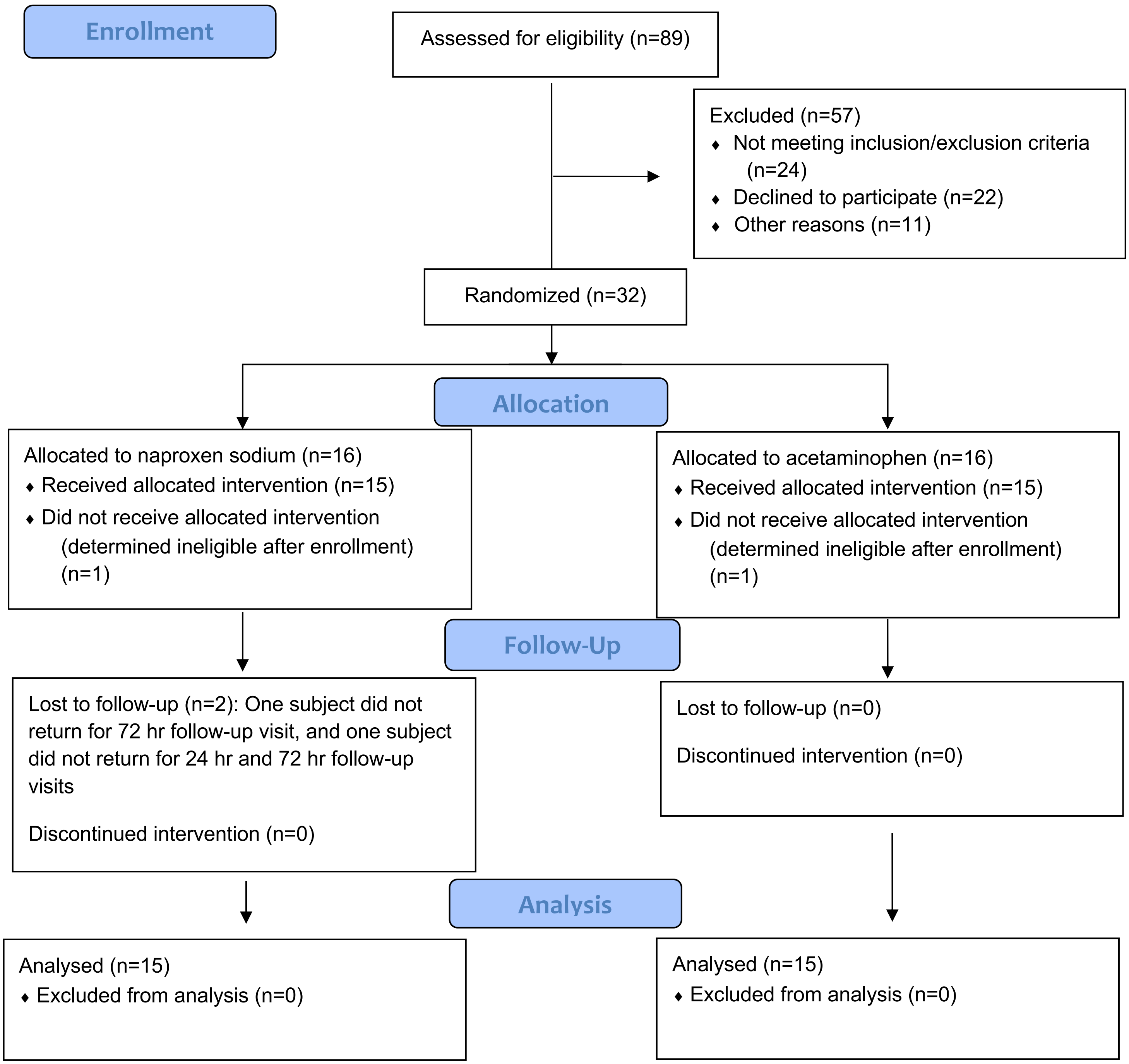
